# Modeling the Transmission Mitigation Impact of Testing for Infectious Diseases

**DOI:** 10.1101/2023.09.22.23295983

**Authors:** Casey Middleton, Daniel B. Larremore

## Abstract

A fundamental question of any program focused on the testing and timely diagnosis of a communicable disease is its effectiveness in reducing transmission. Here, we introduce testing effectiveness (TE)—the fraction by which testing and post-diagnosis isolation reduce transmission at the population scale—and a model that incorporates test specifications and usage, within-host pathogen dynamics, and human behaviors to estimate TE. Using TE to guide recommendations, we show that today’s rapid diagnostics should be used immediately upon symptom onset to control influenza A and respiratory syncytial virus (RSV), but delayed by up to 2d to control omicron-era SARS-CoV-2. Furthermore, while rapid tests are superior to RT-qPCR for control of founder-strain SARS-CoV-2, omicron-era changes in viral kinetics and rapid test sensitivity cause a reversal, with higher TE for RT-qPCR despite longer turnaround times. Finally, we illustrate the model’s flexibility by quantifying tradeoffs in the use of post-diagnosis testing to shorten isolation times.

## Introduction

Four years after the emergence of SARS-CoV-2, a new status quo for test usage has emerged. Despite documented successes of routine screening for SARS-CoV-2 in nursing homes [1], college campuses [2], and even nations [3], regular screening via RT-qPCR or rapid diagnostic tests (RDTs) has given way to elective testing after known exposures or symptom onset, typically with RDTs alone. At the same time, the variety and targets of available diagnostics continues to grow, with numerous available RDTs for respiratory syncytial virus (RSV; [4]) and influenza A [5], new RDTS for SARS-CoV-2 utilizing exhaled aerosols [6], and simultaneous testing for all three viruses via both multiplex RT-qPCR [7] and rapid antigen lateral flow “triple tests” [8]. Rapid diagnostics have proved valuable for a broader set of pathogens too, including HIV [9] and measles [10], as well as *P. falciparum*, with sufficient impact that rapid diagnosis and treatment have even selected for RDT-escape mutations among *P. falciparum* parasites [11].

Direct empirical estimation of the population-scale impact of testing is difficult, and only possible retrospectively, elevating the value of mathematical modeling that can predict it from first principles. Mathematical models estimating the impacts of testing on transmission [2, 12–19] and treatment [20, 21] proved useful in guiding policy and recommendations for SARS-CoV-2, building in many ways on a broad set of earlier efforts to estimate transmission reduction, clinical impact, and cost effectiveness for routine HIV screening (e.g., [22, 23]). However, these successes have been restricted to a limited number of pathogens and either routine screening or risk-group based testing, highlighting the need for more flexible modeling to accommodate an increasing array of diagnostic tests for a growing set of pathogens, used electively after known exposures or symptom onset. Moreover, because testing guidelines are only as effective as human behaviors allow them to be, it would be valuable for models to incorporate key behaviors such as imperfect participation [9] and compliance [24] and imperfect adherence to post-diagnosis isolation [25].

Here, we fill this gap by introducing a more general mathematical model for testing without restricting analysis to a single pathogen, test, testing pattern, or set of behaviors. Our focus is to estimate (i) the extent to which testing reduces the risk of transmission for infected individuals, (ii) the distribution of diagnosis times and the probability that individuals are diagnosed at all, and (iii) the costs of test consumption and isolation days corresponding to these diagnosis and transmission mitigation benefits. Taken together, this model places various intuitions about disease mitigation via testing on firm quantitative ground, and exposes important testing-associated variables and behaviors to *in silico* experimentation and optimization.

To demonstrate how one might use our model, we apply it to the study of the respiratory pathogens RSV, influenza A, and SARS-CoV-2. We first analyze differences between pathogens and between testing strategies by asking whether a single testing strategy might be optimal for all three focal pathogens. Second, we ask when one should test after respiratory symptoms appear, acknowledging that supplies may be limited and infection may not yet be detectable at symptom onset. Third, we reevaluate the tradeoffs between test sensitivity and turnaround time for SARS-CoV-2 in light of shifts in viral kinetics and symptom onset time caused by immune experience and new variants. Finally, we compare the costs and benefits of fixed-duration and test-to-exit isolation guidelines using the model’s cost estimates for test consumption and days spent in isolation. In this way, this paper has twin goals: to first introduce a flexible mathematical model for testing, and then demonstrate its value in the service of relevant scientific and policy questions.

## Results

### A model for testing effectiveness (TE)

To examine the impact of testing on community transmission, we developed a probabilistic model which integrates four key elements: (i) the properties of a particular diagnostic test, (ii) a strategy for its administration, (iii) the time-varying profiles of infectiousness, symptoms, and detectability over the course of an infection, and (iv) the key behaviors of participation (whether or not one refuses to test), compliance (whether or not one takes a recommended test), and isolation length. Given these elements, the model generates a distribution of probable diagnosis times, and uses them to compute the expected reduction in the risk of transmission due to testing.. By then incorporating heterogeneity between individuals, the model estimates *testing effectiveness*, the proportion by which a testing program reduces population-level transmission, in expectation, given by the relationship between reproductive numbers *R* with and without testing,

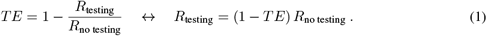

This definition intentionally mirrors the language of vaccine effectiveness (*V E*) against onward transmission, that is, the expected reduction in post-infection transmission risk for vaccinated vs unvaccinated individuals [26, 27]. However, we name the present quantity *testing effectiveness* instead of *test effectiveness* due to the simple observation that the same test, used differently, may have a markedly different impact.

We built our analyses around a simple, common, and computationally efficient model of within-host pathogen kinetics: after some post-exposure latent period, the pathogen load grows exponentially at some proliferation rate until reaching a peak, and then declines exponentially at some clearance rate. This leads to a piecewise linear model of log pathogen load which requires only the four parameters of latent, proliferation, and clearance periods, and a peak load, which we draw from distributions estimated from studies of RSV [28, 29], influenza A [30–32], and SARS-CoV-2 measured for both the founder strain in naive hosts during the prevaccination era [13,17,33] and omicron variants in experienced hosts during the post-vaccination era [33,34] (see Table S1). While the results presented here utilize this simplistic model of viral kinetics, the modeling framework is highly flexible to incorporate more sophisticated alternatives.

We used stochastic realizations from this simple pathogen load model in three ways. First, we assumed that a test taken at time *t* would return a negative diagnosis if the pathogen load was below the test’s limit of detection (LOD), and would return a positive diagnosis with some probability when pathogen load was above the LOD. Due to the importance of test turnaround time [13], we modeled sample-to-answer delays by returning results after a specified turn-around time (TAT). Second, we took infectiousness to be proportional to the logarithm of pathogen load in excess of an empirically estimated threshold [35, 36], consistent with observations that higher viral loads are associated with more efficient transmission for pathogens including SARS [37], SARS-CoV-2 [24], influenza [38], and RSV [39]. Alternative relationships between pathogen load and infectiousness are possible [13, 36]. Third, we drew symptom onset times relative to the times of peak pathogen load, reflective of the typical manner of reporting in the literature [38, 40]. In this way, our model is similar in spirit to the CEPAC model (Cost-Effectiveness of Preventing AIDS Complications), which provides stochastic individual-level realizations of post-HIV-infection dynamics, costs, and outcomes [41].

Our model generates its estimates by integrating over the timing of possible symptoms and tests, to calculate a distribution of diagnosis times, including the possibility of no diagnosis at all. If receiving a diagnosis, each individual is assumed to isolate thereafter for a specified number of days, or until released by a negative test (via a so-called test-to-exit plan), with mitigated infectiousness during isolation (Fig. 1A). By averaging outcomes over the ensemble defined by its random variables, whether by integration or via Monte Carlo, the model produces estimates of the expected infectiousness curves over time, with and without testing (Fig. 1B). The areas under these two curves are proportional to the total transmission potential with and without testing, and thus, their respective reproductive numbers. The model also provides a curve representing the cumulative probability that a randomly chosen infected individual has not yet been diagnosed by some time (Fig. 1B). The long-time limit of this curve is particularly useful because it represents the proportion of the infected population who escape diagnosis entirely. Its complement, the proportion of the infected population to receive a diagnosis at any point, is therefore the ascertainment of the testing scenario, a quantity also called protocol sensitivity in the literature [19]. A complete mathematical description and details of parameterizations can be found in Materials and Methods.

**Figure 1:**
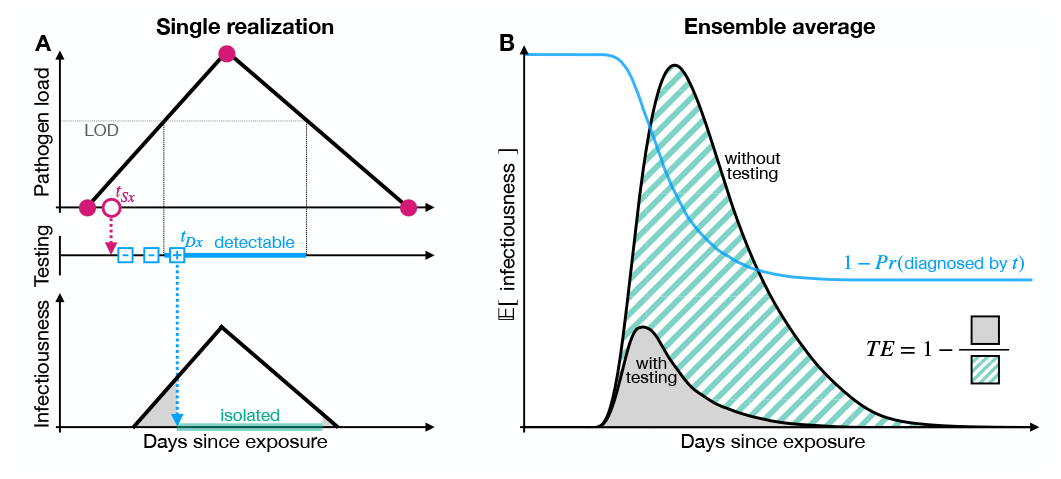
Model diagram. (A) Each realization of the stochastic testing model first draws four control points to specify a piecewise linear model of pathogen load on a logarithmic scale (filled pink circles) and a symptom onset time (*t*_Sx_, open pink circle). The realization then draws a set of testing times (open blue squares), which may be triggered by symptoms (pink arrow), triggered by a known exposure (not shown), or ongoing at a particular cadence (not shown). A test taken during the detectable window (blue bar) when pathogen load exceeds the test’s limit of detection (LOD, gray line) will return a positive diagnosis with a fixed probability after a specified turnaround time (not shown). However, not all tests are necessarily taken, due to imperfect compliance (not shown). Diagnosis at time *t*_Dx_ leads to isolation and thus reduced infectiousness (grey). (B) The ensemble mean, whether computed through integrals or estimated via Monte Carlo, produces expected infectiousness curves with and without testing. The areas under these curves are proportional to their respective reproductive numbers, enabling estimation of testing effectiveness TE (see Eq. (1)). The model also computes the proportion of individuals who remain undiagnosed at time *t* (blue curve), a curve which approaches zero as ascertainment approaches 100%.

### Testing effectiveness varies by strategy and pathogen

Despite the 2023 end to the World Health Organization’s COVID-19 public health emergency [42], the burden of COVID-19 and respiratory viruses more broadly remains substantial. In the U.S. alone, an estimated 9 million cases of influenza A caused 100,000 hospitalizations (2021-2022 season; [43]), and a global estimate of 33 million RSV infections in children under 5y led to 3.6 million associated hospitalizations and 101,400 associated deaths, the vast majority of which were in low- and middle-income countries (2019; [44]). With the broad expansion of diagnostic testing globally, including at-home rapid diagnostic tests (RDTs) for influenza A, RSV, and SARS-CoV-2, we sought to determine whether a single testing strategy might be optimal for all three common respiratory pathogens.

To examine the potential impacts of testing, we considered three distinct testing behaviors, meant to capture both institutional testing strategies and elective testing in response to exposure or symptoms. First, we considered routine weekly RDT screening (turnaround time TAT = 0, LODs Table S1). Given the markedly different sensitivities of different RDT kits and RT-qPCR protocols [45, 46], a representative limit of detection (LOD) was chosen for each respiratory virus to investigate general principles of testing (see Materials and Methods). To incorporate the fact that not all policy-prescribed tests are actually taken in practice [24], this scenario included compliance of only 50%, such that each test was taken or not taken independently with probability 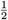. Second, we considered an elective testing scenario in which individuals experiencing symptoms used one RDT per day for 2 consecutive days following the onset of symptoms (TAT = 0; see Table S1 for stochastic timing and prevalence of symptoms). Finally, we considered an elective testing scenario in which 75% of individuals sought out a single RT-qPCR test (TAT = 2, see Table S1 for LODs) between 2d and 7d after exposure; the other 25% did not participate. For each scenario, and each of the three circulating respiratory viruses (RSV, influenza A, and SARS-CoV-2), we calculated TE, the timing of diagnoses, and ascertainment—the total fraction of individuals receiving a positive diagnosis—based on 10^5^ stochastic realizations of viral load, symptom onset, testing, diagnosis, and isolation. For SARS-CoV-2 we specifically considered within-host dynamics and RDTs associated with omicron variants in immune-experienced hosts.

This analysis demonstrated that a test, how it is used, and the pathogen it targets, can interact in potentially complicated ways. For instance, we found that weekly RDT screening with 50% compliance exhibited low TE for all three pathogens, ranging from 13% for omicron-era SARS-CoV-2 down to just 7% for influenza A (Fig. 2A). These low values are driven by the fact that the time windows during which the pathogens are detectable via RDT are almost always shorter than 7d, meaning that weekly testing missed a considerable number of infections (Fig. 3). These short detection windows are compounded by the assumed 50% compliance, leading to low ascertainment across pathogens (11-14%; Supplementary Fig. S1).

**Figure 2:**
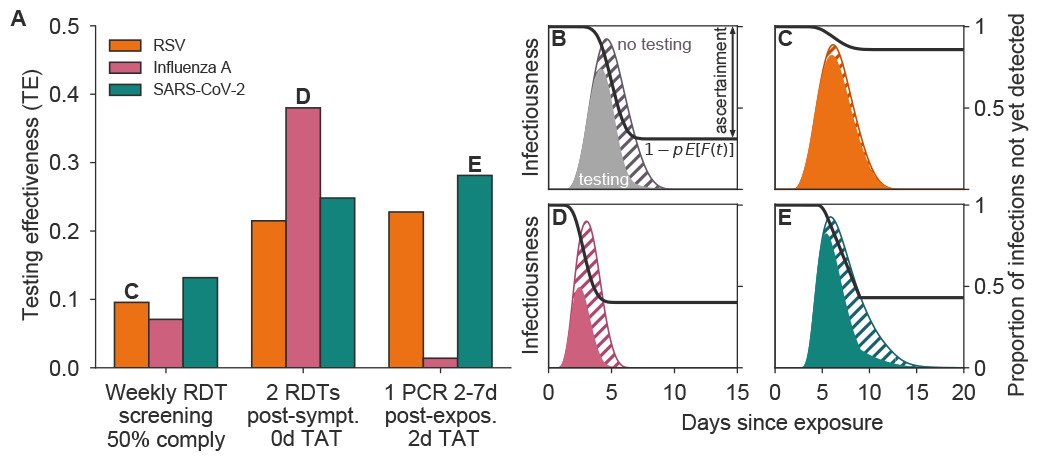
Testing effectiveness varies considerably by strategy and pathogen. (A) Testing effectiveness is shown for RSV (orange), influenza A (pink), and SARS-CoV-2 omicron in experienced hosts (green) under three testing programs: (1) weekly rapid diagnostic test (RDT) screening with 50% compliance, (2) testing with one RDT per day for two days starting at symptom onset, and (3) one RT-qPCR test administered 2-7d after exposure, with 75% participation and 2d test turnaround time (TAT). Panels C-E depict population-level infectiousness curves without (hatched) and with (filled) testing and isolation for the labeled pathogen and testing program, and panel B provides annotations for an example simulation. Black curves represent the proportion of infections not yet detected by time *t*. See Figure S1 for scenario ascertainment rates and Figure S2 for population-level infectiousness curves for all testing scenarios.

**Figure 3:**
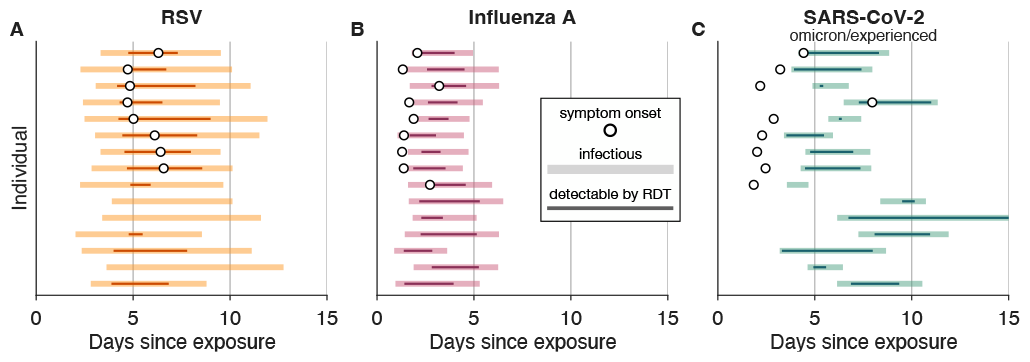
Relative timing of symptom onset, infectiousness, and detectability vary by pathogen and individual. Symptom onset time (open circles), infectious period (shaded rectangles), and window of detectability by a rapid diagnostic test (RDT, colored lines) are shown for 15 stochastic realizations of RSV (A, orange), influenza A (B, pink), and SARS-CoV-2 omicron/experienced (C, green) infections. The absence of an open circle indicates an asymptomatic infection. See Supplementary Table S1 for parameters and references.

In contrast to regular screening, elective testing with the same RDT at symptom onset achieved 38% TE for influenza A, but only 21% for RSV (Fig 2A). This difference in effectiveness primarily reflects differences in the relative timing of symptoms, infectiousness, and detectability: both influenza A and RSV typically become detectable by RDT within one day of symptom onset, but RSV infections are characterized by higher rates of asymptomatic and pre-symptomatic infectiousness (Fig. 3A,B). In contrast, TE for elective testing at SARS-CoV-2 symptom onset was middling at 25%, balancing early detection for those who test positive with lower ascertainment as a result of testing too early to be detected (Fig. 3C). Due to their highly overlapping symptom sets, our results indicate that testing immediately after symptom onset with a single three-pathogen RDT [8] is therefore likely to mitigate transmission most for influenza A, followed by SARS-CoV-2 omicron and RSV.

This ordering of differential TE was inverted for a single elective RT-qPCR between 2d and 7d post-exposure, which was least effective for influenza A (Fig. 2A). This ordering reflects a substantially faster onset of infectiousness after exposure for influenza A (Fig. 3B), meaning that a post-exposure test that is timed effectively for SARS-CoV-2 and RSV is administered far too late to effectively control influenza A. Thus, in a scenario where an individual seeks a highly sensitive multiplex diagnostic test within one week after a known exposure to an unknown respiratory pathogen, our results indicate an impact on transmission of 28% for SARS-CoV-2, 22% for RSV, and just 2% for influenza.

Our results highlight the fact that variation in viral kinetics, symptom onset time, tests’ analytical sensitivities, and interactions thereof, lead to markedly different impacts on transmission, even under the same testing guidance. Given these complexities, we explored whether varying either the pathogen load above which individuals are considered infectious or the RDT limit of detection would substantially alter our findings. These sensitivity analyses showed that, while the magnitude of TE varies slightly, the primary trends documented above remain consistent (Supplementary Fig. S3).

Throughout these experiments, we observed that ascertainment—the proportion of infections diagnosed via testing—was only weakly related to TE, showing that a testing program’s information value and mitigation impact are distinct quantities. For instance, elective RDT testing post-symptoms for influenza A showed TE and ascertainment of 38% and 60%, respectively; for elective RT-qPCR testing post-exposure, TE decreased by 36 percentage points to just 2% but ascertainment decreased by only 10 percentage points to 50% (Supplementary Fig. S1). To illustrate the reason for this difference, we plotted the infectiousness curves

*β*(*t*) with and without testing, averaged over all 10^5^ simulated individuals, as well as the curves showing the fraction of individuals remaining undiagnosed 1 *− p* 𝔼 [*F* (*t*)] (Fig. 2C,D,E). In instances where diagnoses typically arrive earlier, the average *β*(*t*) (and thus the area beneath it, *R*_testing_) is more substantially reduced, while the same number of diagnoses, arriving later, leave more area under the *β*(*t*) curve. Thus, TE incorporates not just whether one is diagnosed, but also when.

### Impacts of timing and availability of elective post-symptom testing

In an era of increasing elective and self-administered rapid diagnostic test (RDT) usage, two key questions are when to test and how many tests to use. We sought to answer these questions by modeling the impact of changes in timing and supply of RDTs on TE for RSV, influenza A, and SARS-CoV-2 omicron in experienced hosts. In these experiments, we considered that individuals would wait 0d-5d after symptom onset and then begin using one RDT per day, with 1-6 RDTs available. For comparison, we also computed TE for a single RT-qPCR with a two-day turnaround. For each fixed supply of tests, we then identified the optimal number of days post-symptoms that one should wait to maximize TE, separately for each pathogen (Fig. 4, white stars).

**Figure 4:**
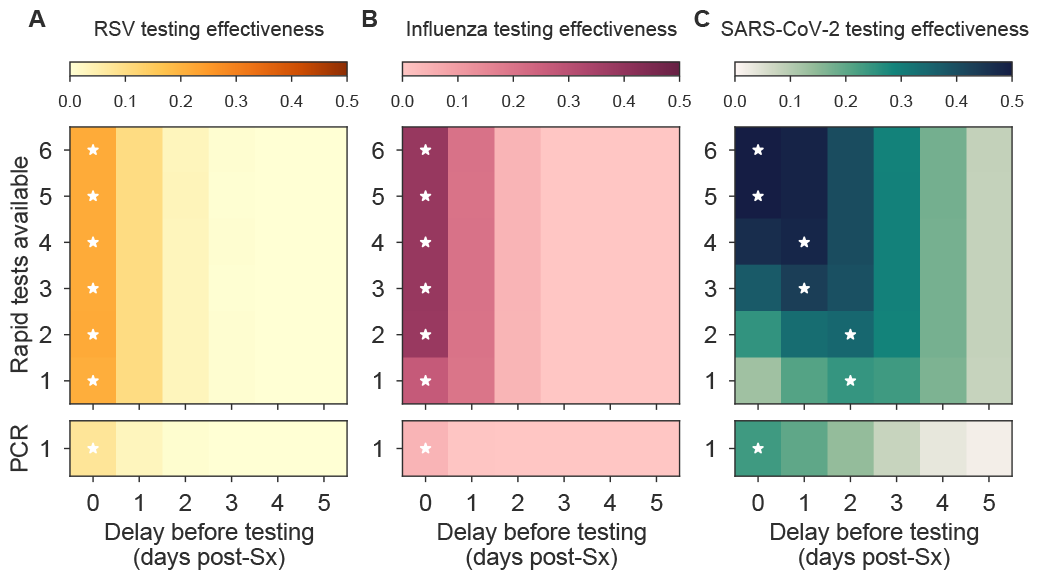
Optimal use of tests depends on the number of tests available and when they are used. Testing effectiveness (TE) of rapid test (RDT) and RT-qPCR with 2 day turnaround time, used *x* days after symptom (Sx) onset using *y* tests once per day is shown for RSV (A, orange), influenza A (B, pink), and SARS-CoV-2 omicron in experienced hosts (C, green). Darker colors represent higher TE as indicated. In each row, the testing strategy with highest TE is annotated with a white star. Turnaround times: rapid tests, TAT = 0; RT-qPCR TAT = 2. See Supplementary Table S1 for LODs and Figures S4 and S5 for monochromatic TE and ascertainment visualizations, respectively.

This experiment showed three common patterns for RSV and influenza A. First, the most effective timing of post-symptom testing was zero days, with TE decreasing monotonically for each additional day of delay (Fig. 4A,B). Second, although using two tests was superior to using one, using more than two tests was roughly equivalent to two. And third, using just one RDT provided superior TE to a single RT-qPCR with a two-day turnaround time, highlighting the importance of RDT availability and rapid results for transmission control.

In contrast, for SARS-CoV-2 omicron in experienced hosts, the number of available tests markedly shifted the optimal time at which one should begin testing, such that when only 1-2 RDTs were available, daily testing was most effective beginning 2d post-symptoms; with 3-4 RDTs, 1d; and with 5-6 RDTs, testing should begin immediately upon symptom onset (Fig. 4C). These results reflect a tradeoff arising from the variable gap between symptom onset and first detectability by RDTs (Fig. 3C): using tests later improves the probability of diagnosis but decreases the impact per diagnosis. A large test supply alleviates this tradeoff by enabling one to test early, with the potential for high impact, while also testing later to avoid missing a diagnosis entirely. Furthermore, for a fixed delay before testing, using more SARS-CoV-2 RDTs was always superior, yet a single RT-qPCR on the first day of symptoms provided approximately equivalent TE to using one RDT starting on day two.

Together, these results suggest a unified recommendation for the timing of post-symptom testing via RT-qPCR for all three pathogens and post-symptom RDTs for RSV and influenza A: one should test immediately, regardless of the number of available tests. In contrast, the timing of optimal SARS-CoV-2 RDT use in supply-limited scenarios depends on the number of available tests. In sensitivity analyses varying the RDT limits of detection and changing the pathogen load threshold for infectiousness, these general findings were unchanged (Supplementary Fig. S6), though the precise recommended post-symptom wait time for SARS-CoV-2 RDTs shifted in some sensitivity analyses by up to 1d.

One common point surfaced by our investigations of elective post-symptom testing was that the existence of any asymptomatic and presymptomatic transmission implies that *TE <* 1 for symptom-driven testing, regardless of the quality of the diagnostic test itself. This implies that even groundbreaking advances in diagnostic LODs, cost, or turnaround times must be paired with appropriate recommendations for usage.

Finally, we briefly note that identical questions of when to test and how many tests to use also arise after known exposure to a pathogen. An otherwise identical analysis recommends wait times of 3-5d after RSV or SARS-CoV-2 exposure, and 0-2d after influenza A exposure, depending on the number and type of available tests (Supplementary Figure S7).

### Reevaluation of the sensitivity/turnaround tradeoff for SARS-CoV-2

Modeling studies in 2020 and 2021 argued that test sensitivity was secondary to frequency and turnaround time for SARS-CoV-2 screening [13,14,47,48], using within-host dynamics and rapid diagnostic test (RDT) sensitivities for the founder strain in naive hosts. However, three important observations regarding the omicron variants circulating in 2023 led us to revisit these findings. First, studies of viral load trajectories, estimated via prospective longitudinal sampling, show lower peak viral loads, shorter clearance times, and slightly longer proliferation times for omicron infections in experienced hosts compared to founder-strain infections in naive hosts [17, 34], leading to shorter windows of detectability (Fig. 5A). Second, symptom onset is typically 3-5d earlier for omicron/experienced vs founder/naive, relative to peak viral load, an observation argued to be due to the immune experience of hosts in particular [34,49,50]. Third, the analytical sensitivity of RDTs is estimated to have worsened for omicron variants vs founder strain, with limits of detection (LODs) increasing by 0.5 *−* 1.0 orders of magnitude depending on the test [45, 46]. Together, these factors led us to hypothesize that the previously established superiority of RDTs over RT-qPCR for mitigation of founder-strain SARS-CoV-2 in a naive population could be equalized or reversed for omicron-variant SARS-CoV-2 in an immune experienced population.

**Figure 5:**
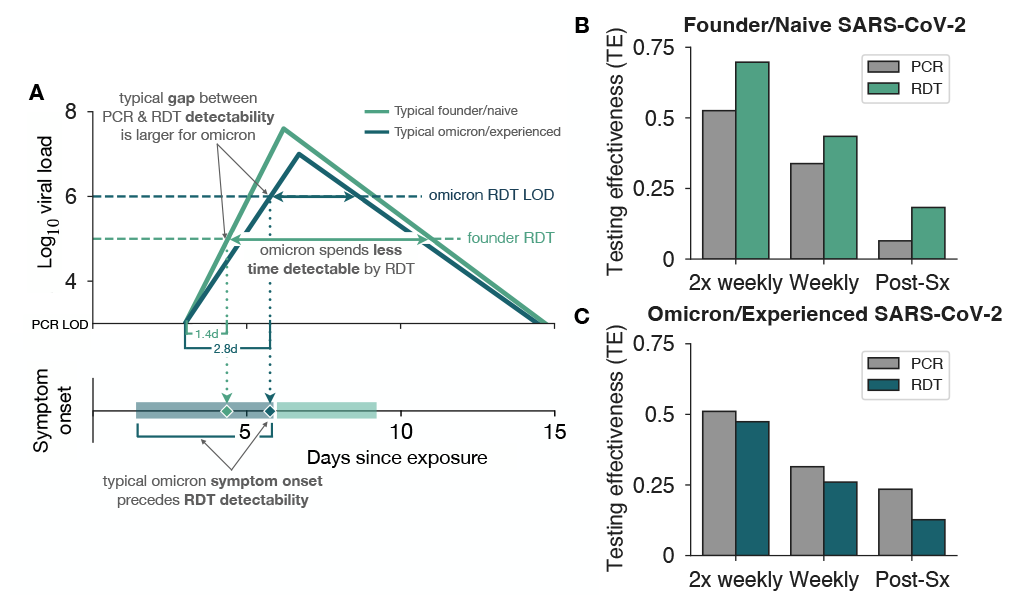
RT-qPCR vs RDT tradeoffs for the SARS-CoV-2 omicron era. (A) Typical viral kinetics for founder SARS-CoV-2 strains in naive hosts and SARS-CoV-2 omicron variants in experienced hosts (log_10_ cp mRNA / mL). Trajectories are characterized above the RT-qPCR limit of detection (LOD = 10^3^) with their respective rapid diagnostic test (RDT) LODs indicated by horizontal dashed lines (10^5^ and 10^6^ for founder strain and omicron variant, respectively). Individuals are considered infectious when the viral load exceeds 10^5.5^ cp mRNA / mL. Potential symptom onset times for each trajectory are shaded on the lower axis. (B,C) Testing effectiveness using RT-qPCR with 2 day turnaround time (gray) or RDT with immediate delivery of results (green) for twice weekly and weekly screening, or testing immediately upon symptom (Sx) onset using one test.

To test our hypothesis, we estimated TE for RDT testing (0d turnaround time, TAT) and RT-qPCR testing (2d TAT) programs, in twice-weekly, weekly, and elective post-symptom testing scenarios, and compared our findings for founder/naive vs omicron/experienced parameters. We found that in each of the three founder/naive testing scenarios, RDTs provided higher TE than RT-qPCR (Fig. 5B), replicating the claims of the literature [13, 14, 47, 48]. However, each of the three omicron/experienced scenarios saw a reversal, with RT-qPCR providing higher TE than RDTs, despite the modeled 2d RT-qPCR turnaround time (Fig. 5C). In general, we also observed that TE decreased for RDTs from the founder-strain era to the omicron era (Fig. 5B vs C), while staying approximately the same (twice weekly, weekly) or even increasing (post-symptom) for RT-qPCR. Together, these results suggest that, setting aside any differences in cost or regulatory complexity, RT-qPCR-based SARS-CoV-2 testing would be superior to otherwise identical RDT-based testing in the omicron and immune-experienced era.

To what can we attribute this apparent reversal in the prioritization of speed vs sensitivity, and how might such principles generalize? First, we note that during viral proliferation, there exists a gap between the time of first detectability via RT-qPCR and the time of first detectability via RDT. This gap represents a potential diagnostic advantage for the RT-qPCR test, but it can be realized only if (i) a test is actually taken during the gap, and (ii) the turnaround time for the RT-qPCR is smaller than the gap. For founder-strain SARS-CoV-2 in the naive host, the typical gap is just 1.4d, vs 2.8d for SARS-CoV-2 omicron variants in experienced hosts (Fig. 5A). Thus, changes in viral proliferation and RDT limits of detection lead to a longer “head start” for RT-qPCR in the era of omicron variants. Second, we note that shortening the window of analytical detectability reduces the chance of diagnosis. For founder-strain SARS-CoV-2 in the naive host, the typical RDT detection window lasts 6.6d, vs 2.2d for SARS-CoV-2 omicron variants in experienced hosts. Thus, RDTs in the omicron variant era have fewer opportunities to contribute to testing effectiveness than those in the founder strain era. We expect both the head start and detection window intuitions to generalize, yet caution that direct estimation of TE is superior to intuition, as not all infections contribute equally to transmission so an apparent advantage for one test over another in the average case may disappear or reverse in the most consequential cases.

### Estimating costs: isolation days and test consumption

Diagnosis-driven isolation provides a mitigation benefit of TE, but at the cost of testing resources and days spent in isolation. Therefore, in addition to quantifying the benefits of TE and ascertainment, we estimated the costs of test consumption and isolation days (Materials and Methods). When calculating these costs, we also recognized that diagnostic tests may be used to determine when one should exit isolation, via so-called test-to-exit (TTE) guidelines [51], potentially increasing test consumption in order to decrease isolation days, with the additional risk of early release from isolation due to a non-analytical test failure. Based on these modeling needs, we modified our estimates of all model outputs to incorporate TTE strategies (Supplementary Text).

To explore the ways in which our model could assist in evaluating complex cost-benefit tradeoffs across scenarios, we considered elective post-symptom testing for SARS-CoV-2 omicron variants in experienced hosts, using either one or two RDTs daily after symptom onset to attempt diagnosis. Diagnosis was followed by either a fixed-duration isolation of 5d, the minimum time recommended by CDC at time of writing [51], or a TTE policy of daily testing using the same RDT, beginning after a minimum of 2d spent in isolation. In each of the 2 *×* 2 scenarios, we computed TE, the average number of tests consumed per infected individual, and the average number of isolation days per diagnosed individual.

At a high level, using two tests to diagnose led to substantially higher TE than using just one (Fig. 6A), mirrored by an increase in per-infection testing cost from 1 to 1.8 tests for the fixed-isolation scenario, and from 1.4 to 2.4 for the TTE scenario (Fig. 6B). Average isolation days per diagnosis were identically 5d for the two-fixed isolation scenarios by definition, and reduced to 3.2-3.3d for TTE (Fig. 6C). Thus, as intended, TTE programs decreased average isolation days without a large impact on TE, driven by the fact that the most common outcome of TTE was release after just 48-72h, while nevertheless maintaining long isolations for those remaining detectable for longer periods (Fig. 6D).

**Figure 6:**
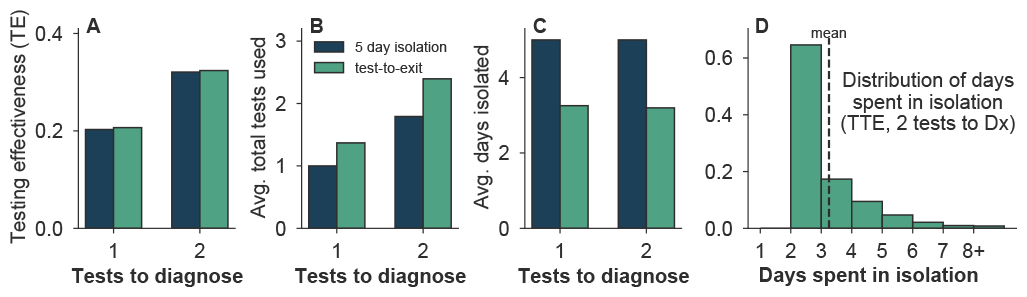
Fixed isolation recommendations may lead to unnecessary isolation when symptomatic testing ascertainment is high. (A) Testing effectiveness, (B) total test consumption over diagnosis and test-to-exit (TTE) usage, and (C) average isolation duration for detected individuals are shown for a fixed 5d isolation period (dark blue) and a test-to-exit isolation program requiring one negative rapid diagnostic test to exit isolation (light green) when 1 or 2 tests were available to diagnose (Dx), used daily beginning one day after symptom onset for SARS-CoV-2 omicron variants in experienced hosts. (D) The distribution of individual days spent in isolation for a test-to-exit scheme using 2 tests to diagnose, with the average isolation time indicated by the vertical dashed line. TTE scenarios assumed individuals waited 2 days after diagnosis before beginning exit-testing.

## Discussion

This study quantifies the impact of testing as a non-pharmaceutical intervention by defining testing effectiveness (TE) as the expected reduction in the risk of transmission for a particular testing behavior, test, and infecting pathogen. While TE varies considerably across the scenarios we explored, three general principles emerge. First, no single elective testing strategy provides superior control across respiratory viruses, due to important differences in their dynamics within host. This highlights the importance of models linking within-host kinetics to between-host transmission when establishing testing guidelines, and the risks of blanket recommendations for respiratory virus transmission control. Second, elective post-symptom or post-exposure testing can have a substantial impact on transmission, but timing is important and depends on the pathogen, test, and available supply. While greater availability of tests leads to strictly larger TE, in supply-limited scenarios, a strategic delay may lead to greater TE by diagnosing more infections, even if they are not detected as early. Last, by modeling tests and testing through different parameters, our analyses show the importance of key behaviors, including compliance, post-diagnosis isolation, and the manner in which to test. These present opportunities to markedly increase the mitigation impact of testing through not only test technology and availability, but through usage guidance and policy as well, based on quantitative guidance from the model.

This work builds on strengths of three established threads in the literature. Many high-quality studies have estimated the value of testing, particularly for routine SARS-CoV-2 screening under a variety of assumptions [2, 12, 14–16, 18–20, 47, 52], and for HIV [9, 22, 23, 41]. Recognizing the growing availability of test options for other pathogens [7, 8, 10] and the collapse of regular SARS-CoV-2 screening programs in favor of elective post-symptom or post-exposure testing, our work proposes a more general framework to meet the current and future realities of testing. This work also builds on the concept of symptom-based controllability [16, 53, 54] by considering a type of diagnosis-driven controllability, and quantifying how much of the theoretical value of symptom-based control can be realized when individuals require a diagnosis before deciding to isolate. Third, this work adds TE to the set of estimates quantifying the effectiveness of targeted interventions that can be directly incorporated as single parameters in between-host transmission models, including vaccine effectiveness [55] and PrEP efficacy [56]. Conceptually, testing effectiveness may be implemented as an increased recovery rate [57] or decreased transmission probability in between-host models, providing both a computational advantage (TE need be estimated only once for a given testing scenario) and the option to incorporate a variable TE as a parameter in other calculations, for instance, to solve for the TE required to achieve *R*_testing_ *<* 1 in a complex compartmental model and then identify testing strategies that achieve it.

By exploring TE under various testing scenarios for three common respiratory viruses, this study demonstrated that an effective strategy for one pathogen may be ineffective for another, and vice versa. This is driven by the complex tradeoffs between a test’s timing, probability of diagnosis, and number of potentially avoided future transmissions should the test come back positive. Intuitively, early testing leads to fewer diagnoses with higher impact per diagnosis, while delayed testing leads to more diagnoses but lower impact per diagnosis. Mathematical models integrating over these contingent factors, stochasticity, and heterogeneity between individual infections, are critical to putting this intuition on a quantitative foundation.

Exemplifying this value, our analysis showed that the use of rapid diagnostic tests upon symptom onset may be an effective strategy for transmission control of RSV and influenza A, but strategic delays in testing may provide higher effectiveness when test availability is limited for SARS-CoV-2. Our analysis also showed that testing with a slower but more sensitive RT-qPCR exhibited a higher TE than otherwise identical rapid antigen testing for SARS-CoV-2 omicron variants in experienced hosts (but not founder SARS-CoV-2 strains in naive hosts), updating a previously published finding that emphasized turnaround time over sensitivity for founder-strain SARS-CoV-2 [13]. Importantly, this analysis focused only on TE under an assumption of identical participation and compliance. However, self-administered rapid diagnostics may have greater availability, lower cost, and higher tolerability than RT-qPCR tests, factors which may lead to differences in participation and compliance which in turn could be incorporated into updated TE estimates. These examples show how TE and its modeling framework may empower researchers or public health strategists to prospectively evaluate diverse testing strategies and behaviors.

The model introduced in this manuscript was necessary because empirical evaluation of the impact of a testing program or behavior is difficult, with compelling analyses nevertheless lacking formal controls [2, 3] or requiring enormous scale [1]. In contrast with various methods to empirically estimate vaccine effectiveness *V E*, we know of no direct identification strategies for testing effectiveness (or similar quantities) in real-world settings. Complicating matters, our calculations suggest that any effective testing program will also shorten the generation interval and increase ascertainment, two quantities that impact the data and models underyling common estimators of the reproductive number *R* [58]. This suggests that an *R*-based empirical TE estimator would present challenges. Methodological development in this direction would be valuable.

Our modeling provides four potentially useful outputs beyond TE. First, we have attempted to quantify the twin costs of test consumption and isolation days, enabling the Pareto frontier between cost and benefit to be explored under a variety of assumptions. When weighted by dollar costs, these estimates may be useful for individuals and policymakers alike. Second, our calculations estimate ascertainment, which may be useful for surveillance and situational awareness. Third, the distribution of *t*_Dx_ *−* TAT, the post-exposure time at which the diagnosing test was taken, could be a valuable inclusion in “nowcasting” models, while the implied distribution of pathogen loads in diagnosing samples may be useful in estimating population-scale epidemiologic dynamics [59]. Finally, in smaller populations, actual testing effectiveness may differ from TE estimates due to finite size effects, variation which could be estimated from sets of Monte Carlo approximations, enabling the incorporation of uncertainty into testing strategy evaluation.

Our work is subject to a number of important limitations associated with the structure of our model. First, we assumed that symptoms may trigger testing, but do not trigger isolation in and of themselves. Relaxation of this assumption would require empirical estimates of self-isolation behavior or simply the additional assumption that a proportion of individuals isolate at symptom onset without testing [13]. Second, we modeled participation/refusal, compliance, and isolation behaviors as statistically independent between individuals, but health-related behaviors are known to be clustered [60–62], a type of heterogeneity not included in our TE calculations. In principle, estimates of TE for a set of behaviorally homogeneous groups could be computed and integrated into appropriately structured transmission models [57]. Finally, our model includes no treatment of specificity, an important factor for certain classes of diagnostic tests. The inclusion of imperfect specificity in our model would not affect TE estimates, but could substantially affect cost estimates, particularly if a low positive predictive value led to markedly more isolation days. For such scenarios, our model could be easily extended to include a second confirmatory test to derive a new distribution for *t*_Dx_.

Our work is also subject to limitations associated with parameterization. For instance, we relied on estimates of pathogen load dynamics and symptom onset time, including variation thereof across a population. These are relatively well characterized for variants of SARS-CoV-2 in moderate to large population cohorts [17, 33,35,36,63], but are sparse for RSV [28,29] and influenza A [30–32] where they come typically from small numbers of healthy volunteers in human challenge studies and are typically presented through population means and confidence intervals, which, at best, indirectly inform between-host variation [64]. Even weakly characterized distributions are effectively unknown for many other pathogens, particularly prior to symptom onset. Joint estimates of viral kinetics and symptom prevalence and timing for many communicable diseases would be powerful for our modeling, and useful in many other applications as well, including studies of the value of early diagnosis as a path to timely treatment [20, 21]. Another important limitation is the assumption that infectiousness can be parameterized as a function of the logarithm of viral load. While viral load and infectiousness are empirically linked in some studies [24, 36, 65], their precise relationship is more complicated. For instance, studies of SARS-COV-2 have shown fewer plaque-forming units per copy of viral RNA during an infection’s clearance phase than during its proliferation phase [50].

Finally, as with any public health intervention, there are important ethical considerations related to this work. This study focused on using resources (tests and isolation days) to provide a benefit (decreased transmission), yet not all individuals, communities, or settings may be equally well positioned to afford tests or isolation time. Similarly, our modeling focuses on population-scale transmission, but ignores heterogeneity in vulnerability among those to whom a disease may be transmitted. Finally, stigma around infection status could lead to low participation, particularly if that otherwise private status will be disclosed [66]. Application of this work should consider affordability, incentives, vulnerability, and privacy in local contexts.

## Materials and Methods

### Mathematical Model for Testing

We introduce testing effectiveness TE as the proportion by which a testing program decreases the risk of transmission, given infection, for an infectious disease [Eq. (1)] and define a mathematical model to prospectively estimate it. This model combines known or assumed properties of (i) a particular diagnostic test, (ii) a strategy for its administration, (iii) behavior/isolation after diagnosis, and (iv) the time-varying profiles of infectiousness and detectability over the course of an infection. Due to its integration of these elements, the model can also estimate a testing program’s ascertainment (the proportion of infections detected), the impact of testing on the generation interval and selection coefficients, the distribution of diagnosis times, and the expected number of tests and isolation days required per diagnosis.

#### Infectiousness

In the absence of testing, the individual reproductive number *ν*_0_ quantifies the expected number of secondary infections from that person [67], under typical behavior. It is given by the area under that individual’s infectiousness curve *β*(*t*) over time,

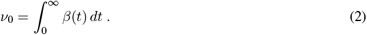

The mean across all individual reproductive numbers is *R* by definition, and therefore

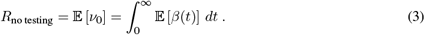

By prompting a post-diagnosis behavior change, participation in testing may isolate or attenuate part of that individual’s infectiousness, decreasing its total from *ν*_0_ to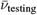. Provided that one’s *ν*_0_ and one’s testing behaviors are statistically independent, then the mean across all individuals’ 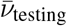 is related to the effective reproductive number as

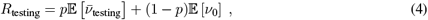

where *p* represents the proportion of the population participating in testing. The participation rate incorporates both the proportion of the population that “opts in” to testing, as well as the fact that even among those opting in, not all will experience symptoms (for elective post-symptom testing) or be alerted to their having been exposed (for elective post-exposure testing). Our focus in the derivation that follows is to estimate 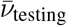 for each person, after which we may average to get *R*_testing_ and thus TE.

#### Isolation after diagnosis

Suppose that at time *t*_Dx_, an individual receives a diagnosis that causes them to attenuate their infectiousness via a change in behavior, such as isolation, masking, or increased ventilation. Here, we develop the mathematics corresponding to perfect isolation post-diagnosis, but provide equations for partial and/or time-varying impacts of behavior on transmission in Supplementary Materials. Post-diagnosis isolation leads to

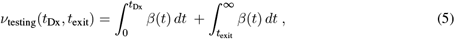

which has the simple interpretation that one’s infectiousness is simply that which occurred prior to diagnosis at *t*_Dx_ and that which occurred after exiting isolation at *t*_exit_. While more complicated test-to-exit equations are developed in Supplementary Materials, here we consider only fixed isolation periods of duration *l*, such that *t*_exit_ = *t*_Dx_ + *l*.

#### Time of diagnosis as a random variable

In practice, the time of diagnosis *t*_Dx_ depends on numerous factors including test availability or schedule or the timing of symptoms. We model *t*_Dx_ as a random variable with probability density function *f* (*t*_Dx_). This leads to an effective infectiousness, calculated in expectation over the probable times of diagnosis, of

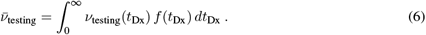

Substituting in the definition of *ν*_testing_(*t*_Dx_) from Eq. (S1), rearranging, and making use of the cumulative probability of a diagnosis *F* (*t*),

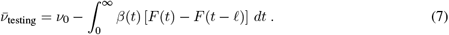

We note that if one reinterprets *F* (*t*) as the cumulative probability of symptom onset (as if symptoms were the diagnostic test itself), with an exhaustive isolation thereafter (large *l*), then Eq. (7) recovers the core notion of symptom-based controllability of Fraser et al. [53]. However, unlike symptom onset, the time of diagnosis via testing *t*_Dx_ depends on within-host kinetics, test administration, and of course the test itself, as we calculate next.

#### Calculating t_Dx_: testing, compliance, failure rates, detectability, and turnaround time

To calculate the distribution of diagnosis times *f* (*t*_Dx_), we combine test administration—the probability that a test is administered at a particular time—and detection—the conditional probability that said test would return a positive diagnosis. This approach naturally separates a test administration strategy from the properties and performance of the particular diagnostic to be modeled.

Let *A*(*t*) be the prescribed rate of test administration in tests per day, such that for sufficiently small *δ*,

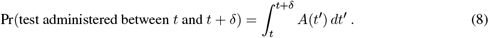

For instance, setting 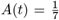 would model a weekly screening regimen, while *A*(*t*) = 2 for 4 *≤ t ≤* 6 would represent twice-daily testing starting 4 days post-exposure and ending on day 6.

Let *D*(*t*) be an indicator function representing the analytical detectability of the infection at time *t* for a given diagnostic, such that *D*(*t*) = 1 when an infection is, in principle, detectable by that diagnostic, and *D*(*t*) = 0 otherwise. For example, to model qPCR, *D*(*t*) = 1 when concentrations of RNA or DNA from a biospecimen exceed the assay’s limit of detection; below the limit of detection, *D*(*t*) = 0.

Combining administration with detectability, the cumulative number of scheduled tests with the potential to return a positive result by time *t* is

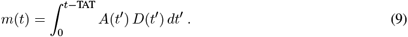

Note that the integral’s upper limit is shifted by TAT, the test turnaround time, i.e., the amount of time between a test’s administration and the return of actionable results. Although any particular person may have taken only an integer number of tests, *m*(*t*) computes a real-valued expectation over all testing schedules or phases that match the specified rate *A*(*t*).

The function *A*(*t*) represents an *intended* test administration strategy (e.g., a policy or guideline), yet in many circumstances, imperfect compliance may result in missed tests. To account for compliance, we let *c* be the independent Bernoulli probability that each test is actually taken as intended.

Similarly, a test may fail for reasons unrelated to the analytical limit of detection or presence of a symptom. For example, poor biospecimen collection technique (e.g. poor nasal swab technique) could result in test failure irrespective of an assay’s limit of detection [45]. To account for test failure of this type, we let *ϕ* be the independent Bernoulli probability that a test fails for non-analytical reasons.

Combining the number of possible positive tests taken *m*(*t*) with compliance *c* and test failure *ϕ*, allows us to compute the cumulative probability that one has received a positive test by time *t*,

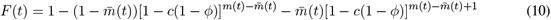

where 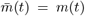 mod 1. This function reflects the cumulative probability that for a monotonically increasing number of possible positive tests *m*(*t*), there are two possibilities: one may have taken either 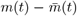 tests with probability 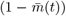, or 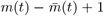 tests with probability 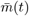. In turn, each of these integer number possible positive tests tests may fail (with probability *ϕ*) or be missed due to non-compliance (with probability 1 *− c*). We note that this is an improper CDF which need not reach 1 in the limit of large *t*, as not all individuals will necessarily receive a diagnosis.

In the case where the product *A*(*t*)*D*(*t*) is a constant *Ā* between *t*_1_ and *t*_2_, and 0 otherwise, the associated improper PDF is

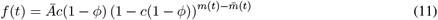

for *t*_1_ + TAT *≤ t ≤ t*_2_ + TAT and 0 otherwise.

In the specific case of elective testing at symptom onset, the functions above require slight modification to include the symptom onset time *t*_Sx_. We modeled *t*_Sx_ as a random offset from the time of peak viral load, drawn from a specified uniform distribution (see Supplementary Table S1), a choice that reflects the way symptom onset is typically reported in the literature. For the cases considered in this manuscript, individuals wait *x* days after symptoms and test for *y* days at rate Ā per day, leading to a test administration function,

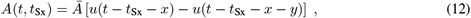

where *u*(*t*) is the unit step function defined as 0 prior to *t* and 1 thereafter. This equation means that Eq. (9) depends on *t*_Sx_, and thus so does the cumulative probability of detection,

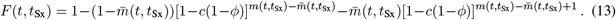

The cumulative probability of detection can then be computed by integrating Eq. (13) against the *t*_Sx_ distribution.

### Model Estimates

#### Testing effectiveness

Testing effectiveness TE can now be computed by substituting Eq. (7) 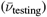 into Eq. (4) (*R*_testing_) into Eq. (1), to get

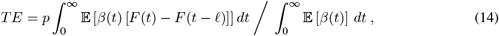

where Eq. (10) can be used to compute the CDF *F* (*t*) of diagnosis times *t*_Dx_. If isolation is of sufficient duration to prevent all post-diagnosis transmission, *TE* simplifies to

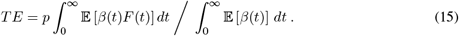

Note also that the integrands enclosed in expectations above have useful interpretations: they represent the expected infectiousness trajectories with (numerator) and without (denominator) participation in testing.

#### Ascertainment, and diagnosis and swab time distributions

For a single individual, the distribution of diagnosis times is given by its improper *f* (*t*_Dx_) or CDF *F* (*t*). The related improper distribution of “diagnosing swab” times is given by shifting the CDF by the turnaround time TAT.

The long-time limit of *F* (*t*) is the probability that that individual will be diagnosed at all. Ascertainment, the proportion of infections diagnosed by a particular strategy at the population scale, can therefore be estimated by taking the expectation of the long-time limits over individuals, and scaling by the participation rate,

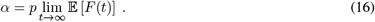

Here, the expectation is taken over individual heterogeneity in within-host dynamics across the population (just as in Eqs. (3) and (4)).

#### Mean generation interval and selection coefficient

The mean generation interval, defined as the typical time between infection and subsequent transmission, can be computed from the centers of mass of the expected infectiousness curves that appear in the numerator and denominator of Eq. (S3). Such estimates may be important for post-hoc estimation of TE from empirical data, given that methods of estimating *R*_*t*_ from empirical case counts often rely on the mean generation interval.

If a single test is capable of diagnosing two strains of the same pathogen, then there is the potential for testing to alter the selection coefficient. Here, TE calculations can estimate this impact, such that the selection coefficient is predicted to shift from 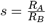 to 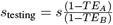 .

#### Test Consumption

How many tests are consumed during the course of a testing regimen? We break this calculation into two cases, depending on whether the individual receives a diagnosis or not.

First, suppose an individual never tests positive. This implies that any tests that were meant to be taken when the infection was detectable, i.e. when *D*(*t*) = 1, were either (i) not taken, due to a failure of compliance (probability 1 *− c*), or (ii) taken yet failed to provide the correct positive diagnosis (probability *cϕ*). Because only the latter case consumes a test, this means that a key quantity is *cϕ/*(1 *− c* + *cϕ*), the relative probability that a planned test consumed a test, given the absence of diagnosis. On the other hand, any tests meant to be taken when the infection was undetectable, i.e. when *D*(*t*) = 0, were taken with probability *c*. Summing these two provides an estimate of average test consumption for those who are not diagnosed.

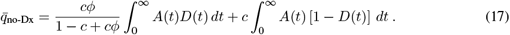

The reason that we call this an estimate and not an exact calculation is that testing and diagnosis are causally linked, and therefore conditioning on a no-diagnosis outcome means that the prescribed test administration may no longer be *A*(*t*) exactly.

In contrast, if an individual receives a diagnosis at *t*_Dx_, we assume that no additional tests were consumed thereafter, and that at least one test was consumed—if not, no diagnosis could have been produced. For a fixed time of diagnosis,

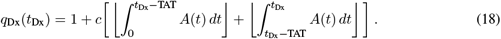

Above, the first integral accounts for non-diagnostic test consumption up until the diagnosing test, while the second integral accounts for additional tests consumed while waiting for the diagnosing test. The separate floor functions are a necessary consequence of conditioning on the separate counting of the diagnosing test. Taking an expectation over *t*_Dx_ yields

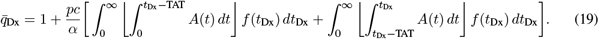

Combining our calculations for 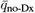 and 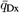, weighting the latter by ascertainment and the former by its complement, we get a general expression for the expected test consumption among infected individuals,

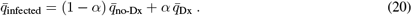

Of course, the whole point of testing is that one does not know *a priori* who is infected and who is not. Consequently, we can estimate consumption by those who are not infected as

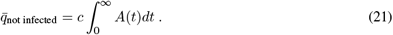

Finally, combining Eqs. (20) and (21), we get

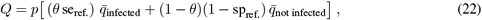

where *p* is the population participation rate, *θ* is the prevalence of the pathogen, and se_ref._ and sp_ref._ are the sensitivity and specificity of the scheme used to refer people to testing, respectively. Such referral schemes include contact tracing, the appearance of symptoms, membership in a group with known high risk of infection, or even universal testing (in which case the referral “program” would have se = 1 and sp = 0).

#### Days spent in isolation

For fixed-duration perfect isolation of length *l*, the average number of isolation days per infected person is given by the product of *l* and ascertainment *d* = *lα*. A similar calculation for test-to-exit policies is found in Supplementary Equation (S10).

### Parameterization of the Model

#### Viral kinetics, symptoms onset, and infectiousness

We parameterized the time-varying profiles of viral load and infectiousness using a simplified model which captures four features common to RSV [28], influenza A [30], and multiple variants of SARS-CoV-2 [17]: (i) a post-exposure period where virus is undetectable by any known test, (ii) a proliferation period of exponential growth, (iii) a peak viral load (VL), followed by (iv) a clearance period of exponential decline. We capture these features using a piecewise linear function describing the logarithm of viral load, specified by three points: (*t*_0_,LLOQ), the first time at which VL exceeds the lower limit of quantification (LLOQ); (*t*_peak_,*V*_peak_), peak VL timing and concentration; and (*t*_*f*_,LLOQ), the last detectable time [13, 17], where *t* represents the time since exposure in days (Supplementary Fig. S8). For the pathogens considered here, the LLOQ is equal to the RT-qPCR LOD. Because within-host kinetics vary from one infection to another, each trajectory is parameterized using independent draws from random variables for each control point.

While summarized in Supplementary Table S1, we briefly review the studies and sources of evidence used to parameterize our simple viral load models. Influenza A latent period parameters were drawn from a review of challenge studies by Carrat et al. supporting a 0.5–2d delay between inoculation and first detectable instance using a gold standard test [30], noting that this range is slightly wider than in other studies [31]. Data from a household study from Ip et al. characterized peak VL between 1–3d after symptom onset, and noted that symptom onset and first detectability were indistinguishable. Their data also showed peak VL between 6–8.5 log_10_ cp RNA/mL, followed by 2–3d clearance times [32]. We note that these clearance times were shorter than those observed in challenge studies [30, 31]. Symptom onset time was specified as taking place between 2d and 0d prior to peak VL from observations of naturally acquired infection [38].

RSV viral kinetics were characterized using vaccine efficacy challenge studies, which reported only geometric means of viral load, measured in days since challenge inoculation [28, 29]. Placebo group data from both studies support a 2–4d latent period and 3-6d between peak VL and clearance [28, 29]. Schmoele-Thoma et al. present individual data points for longitudinal sampling of infected participants supporting a proliferation phase of 2–4d and peak VL between 4.5–8 log_10_ cp RNA/mL [28]. Sadoff et al. indicate slightly later and lower peaks, but we weight these less heavily in model parameterization because only mean and confidence intervals are presented, but not individual data points [29]. Symptom onset time was specified as taking place between 1d prior to and 1d after peak VL from an additional human challenge study [40].

We briefly note that our characterization of influenza A and RSV viral kinetics relies primarily on studies meant to capture vaccine [28, 29] and drug [31] efficacy, or to report symptom dynamics [32]. In all four studies, viral kinetics data are reported as a secondary finding, and typically as a geometric mean since time of onset—including, in two instances, all the individuals for whom inoculation failed [28, 29]. Due to the lack of viral load data linked at the level of individuals to inform distributional choices of kinetics parameters across a population, we assumed uniform distributions over supported parameter ranges for both viruses.

In contrast, our estimates of SARS-CoV-2 viral kinetics parameters were drawn from studies performed specifically to characterize within-host viral dynamics. Kissler et al. and Hay et al. provide mean and 95% credible interval estimates for peak VL and the durations of the proliferation and clearance phases for founder-strain/naive and omicron-strain/experienced SARS-CoV-2, respectively [17, 34]. Parameter distributions were assumed to be Lognormal with parameters *μ* and *σ*, whose values were adapted from published means, 95% credible intervals, and sample sizes. Specifically, we selected values such that an equal number of draws from Lognormal(*μ, σ*) would lead to a frequentist confidence interval matching the published mean and credible interval to the closest approximation. See Table S1 for these parameter values. Given the possibility of the occasional non-biologically large draw from the Lognormal distributions, we rejected proliferation phases shorter than 0.5d and longer than 10d, and rejected clearance phases shorter than 0.5d and longer than 25d for both SARS-CoV-2 models. A challenge study provides support for a 2.5-3.5d latent period after inoculation [33]. Symptom onset time was specified as taking place between 0-3d after peak VL for founder-strain/naive infections [34, 49, 50], and between 1-5d before peak VL for omicron-variant/experienced infections [33, 68, 69].

Given a stochastic realization of viral kinetics from the model above, we calculated infectiousness *β*(*t*) as proportional to the logarithm of viral load in excess of some minimum threshold, specified by the typical viral load (concentration of RNA cpRNA/ml) at which plaque forming units are consistently found (*>* 1 PFU/ml). While this type of log-viral-load infectiousness assumption is common for studies of influenza A [30, 70], SARS-CoV-2 [52], and RSV [71], alternative relationships between viral load (or other quantities) and infectiousness are possible [13, 72].

#### Analytical sensitivity and failure rate of diagnostic tests

Analytical sensitivities (limits of detection; LODs) were drawn from the literature for RT-qPCR and RDT tests for influenza A, RSV, founder-strain SARS-CoV-2, and SARS-CoV-2 omicron variants. Due to the variability in LODs between assays of the same type for the same pathogen, we chose a single value (Table S1 to represent each pathogen and test type. Above a test’s estimated LOD, false negative rates *ϕ* were available only for SARS-CoV-2 [45] at approximately 5%, a rate we assumed for RSV and influenza A, collectively modeling factors such as sample contamination, poor biospecimen collection, or manufacturing error. In general, parameters were more widely available and better characterized for SARS-CoV-2 tests than for RSV or influenza A. See Table S1 for LODs, failure rates, and relevant sources.

## Data Availability

No datasets were generated or analysed during the current study. All code needed to evaluate the conclusions in the paper are present in the paper and/or the Supplementary Materials, and open-source code (Python 3.7.4) is available at https://github.com/CaseyMiddleton/TestingFramework.

https://github.com/CaseyMiddleton/TestingEffectiveness

## Acknowledgements

We thank the BioFrontiers Institute IT HPC group, Yonatan Grad, Sarah Cobey, and Stephen Kissler.

## Author Contributions

C.E.M. and D.B.L. conceived of and designed the study, derived the model, and wrote the manuscript. C.E.M. conducted extensive literature reviews to parameterize models, and conducted numerical simulations.

## Data and Materials Availability

All open-source code (Python 3.7.4) needed to evaluate the conclusions in the paper is available at https://zenodo.org/doi/10.5281/zenodo.10457041. All other data is included in the paper and/or Supplementary Materials.

## Supplemental Figures and Tables

**Figure S1:**
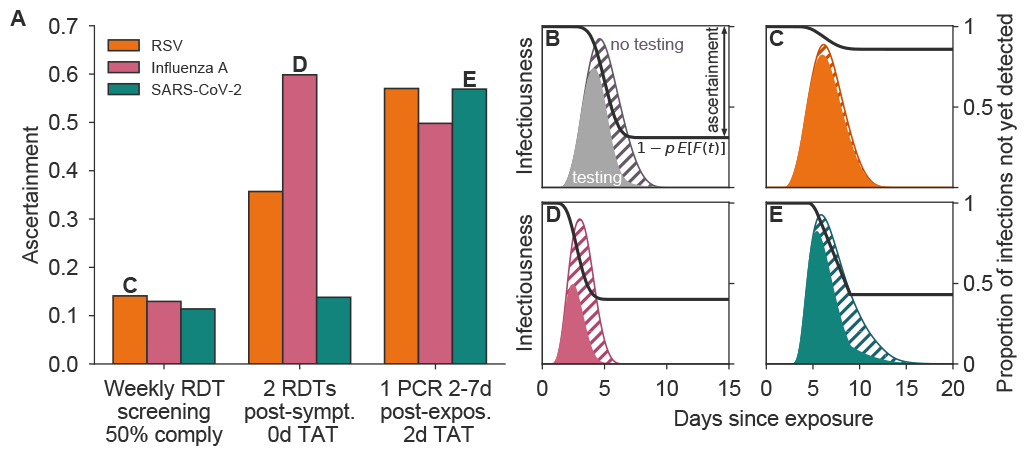
Ascertainment rates vary considerably by strategy and pathogen. Ascertainment is shown for RSV (orange), influenza A (pink), and SARS-CoV-2 omicron in experienced hosts (green) under three testing programs: (1) weekly rapid diagnostic test (RDT) screening with 50% compliance, (2) testing with one RDT per day for two days starting at symptom onset, and (3) one RT-qPCR test administered 2-7d after exposure, with 75% participation and 2d test turnaround time (TAT). Panels B-E depict population-level infectiousness curves without (hatched) and with (filled) testing and isolation for the labeled pathogen and testing program. Black curves represent the proportion of infections not yet detected by time *t*. See Supplemental Figure 2 for scenario Testing effectiveness estimates.

**Figure S2:**
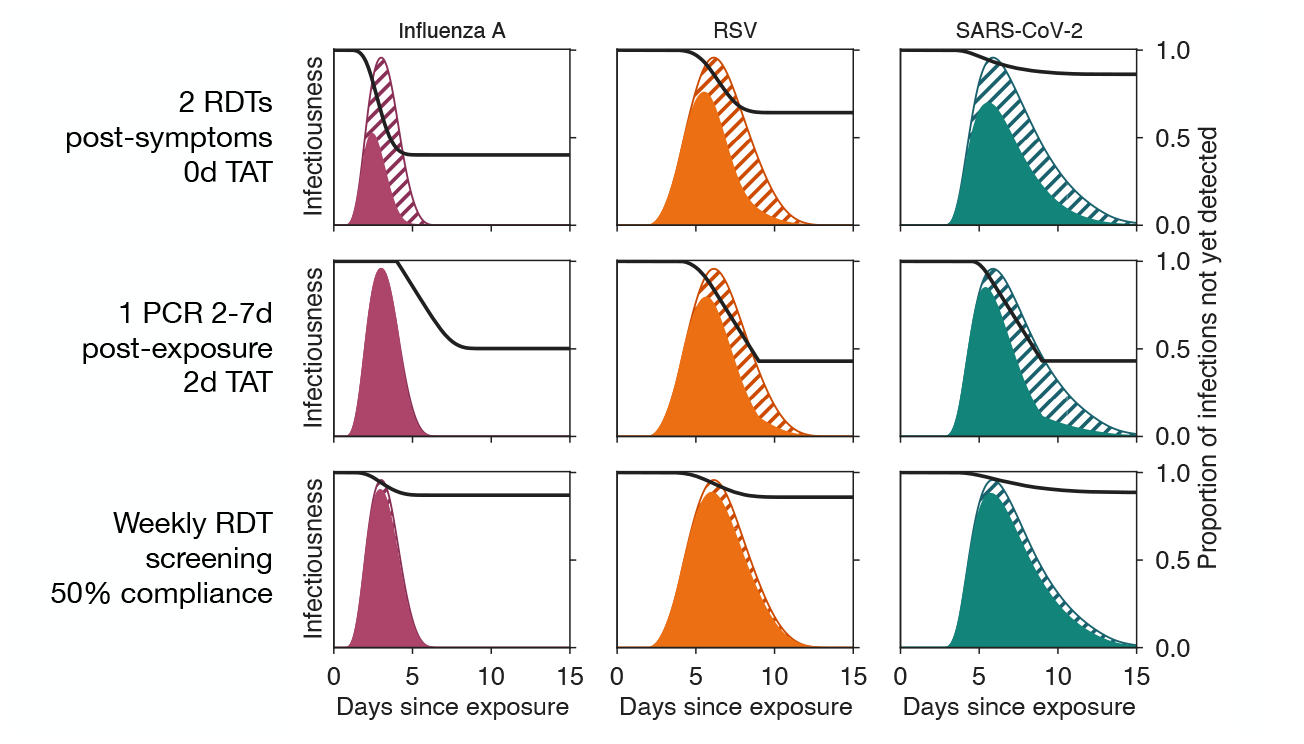
Population-level infectiousness curves for all Figure 2 scenarios. Scenario results are shown for RSV (orange), influenza A (pink), and SARS-CoV-2 omicron in experienced hosts (green) under three testing programs: (1) weekly rapid diagnostic test (RDT) screening with 50% compliance, (2) testing with one RDT per day for two days starting at symptom onset, and (3) one RT-qPCR test administered 2-7d after exposure, with 75% participation and 2d test turnaround time (TAT). Panels depict population-level infectiousness curves without (hatched) and with (filled) testing and isolation for the labeled pathogen and testing program. Black curves represent the proportion of infections not yet detected by time *t*.

**Figure S3:**
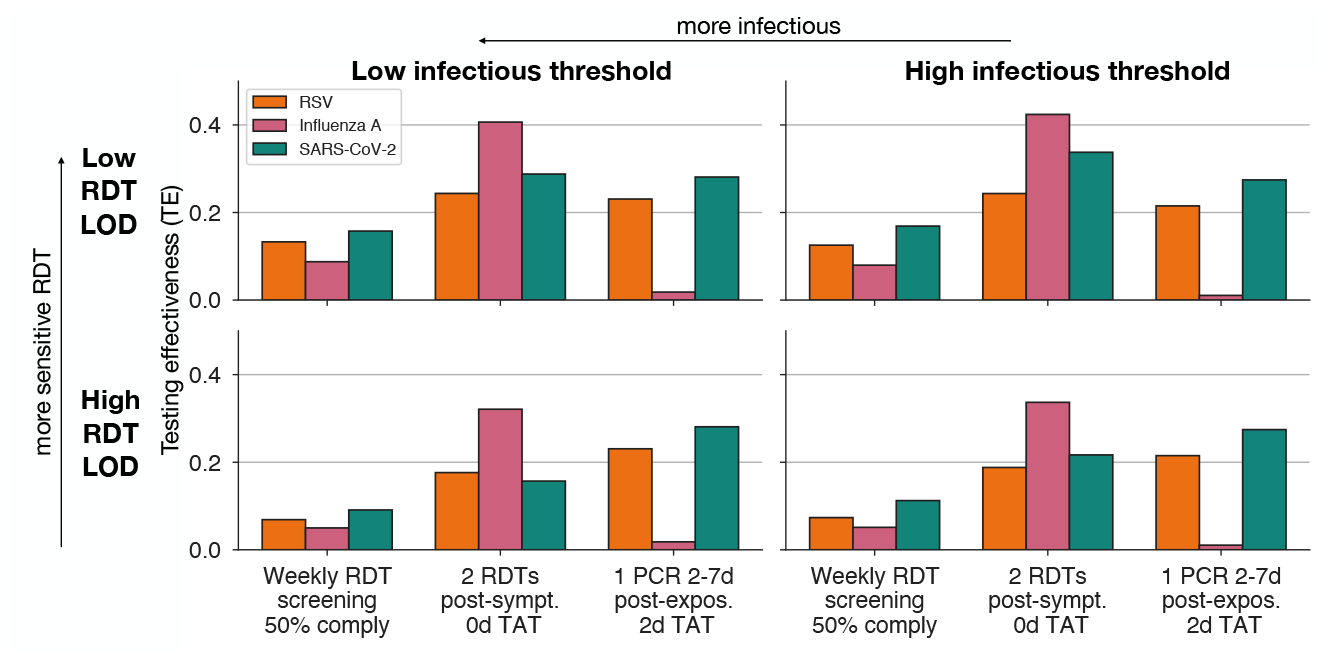
Sensitivity analysis of testing effectiveness patterns observed in Figure 2. Testing effectiveness is shown for RSV (orange), influenza A (pink), and SARS-CoV-2 omicron in experienced hosts (green) under three testing programs: (1) weekly rapid diagnostic test (RDT) screening with 50% compliance, (2) testing with one RDT per day for two days starting at symptom onset, and (3) one RT-qPCR test administered 2-7d after exposure, with 75% participation and 2d test turnaround time (TAT). Each scenario is considered for a 0.5 log-fold increase or decrease in infectious threshold or RDT limit of detection (LOD), as labeled. In this sensitivity analysis, although precise values change, the relative patterns and orderings discussed in the main text are unaffected.

**Figure S4:**
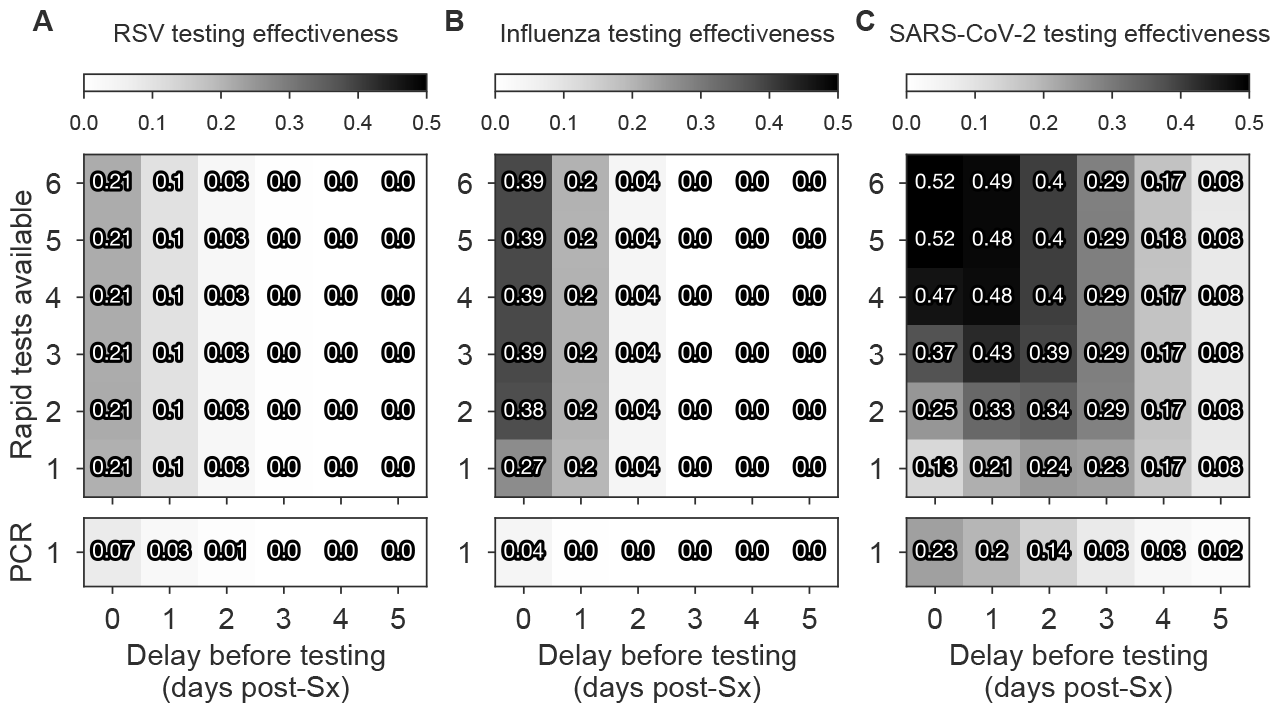
Testing effectiveness after symptom onset depends on the pathogen, the number and type of tests available, and when they are used. Testing effectiveness (TE) of rapid test (RDT) and RT-qPCR with 2 day turnaround time, used *x* days after symptom (Sx) onset using *y* tests once per day is shown for RSV, influenza type A, and SARS-CoV-2 omicron in experienced hosts. Darker colors represent higher TE as indicated, and the associated TE for each strategy is shown. Turnaround times: rapid tests, TAT = 0; RT-qPCR TAT = 2. See Supplementary Table S1 for limits of detection and other parameters and Figure 4 for colored heatmaps with annotated optima.

**Figure S5:**
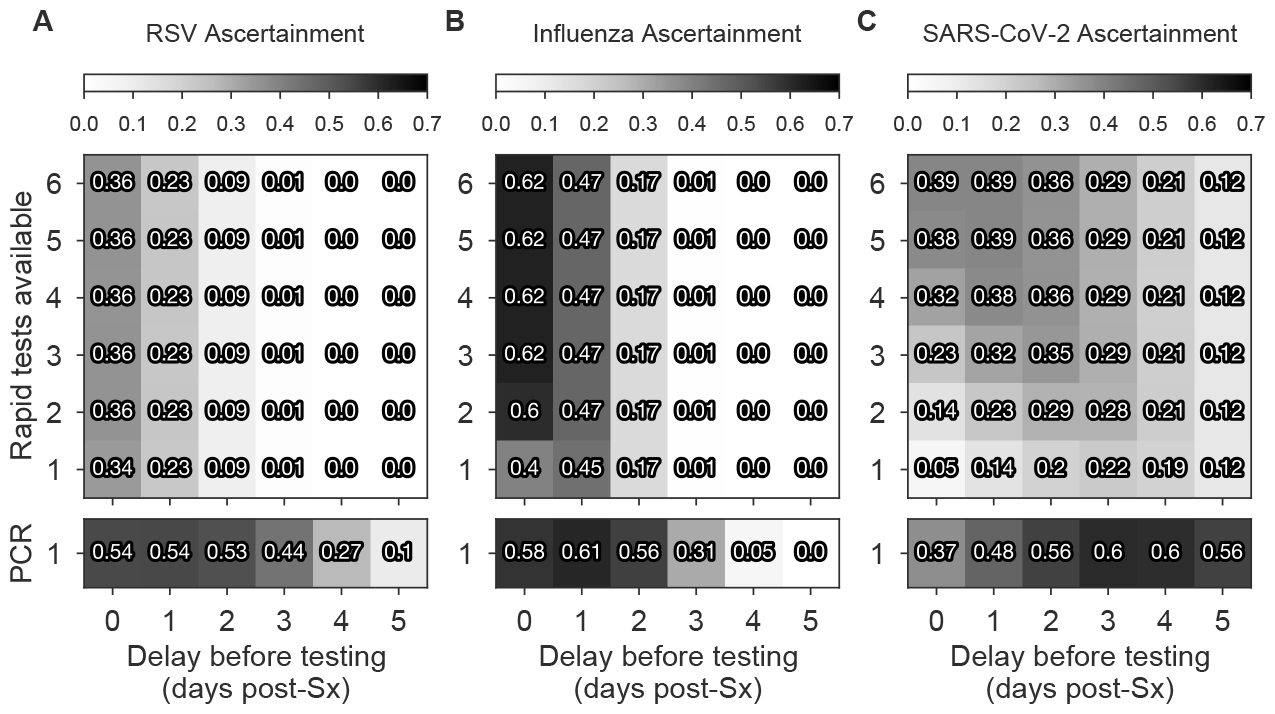
Ascertainment of testing after symptom onset depends on the pathogen, the number and type of tests available, and when they are used. Ascertainment—defined as the total proportion of infections diagnosed—of rapid diagnostic test (RDT) and RT-qPCR with 2 day turnaround time, used *x* days after symptom (Sx) onset using *y* tests once per day is shown for RSV, influenza A, and SARS-CoV-2 omicron in experienced hosts. Darker colors represent higher ascertainment as indicated, and the ascertainment values are shown. Turnaround times: rapid tests, TAT = 0; RT-qPCR TAT = 2. See Supplementary Table S1 for limits of detection and other parameters.

**Figure S6:**
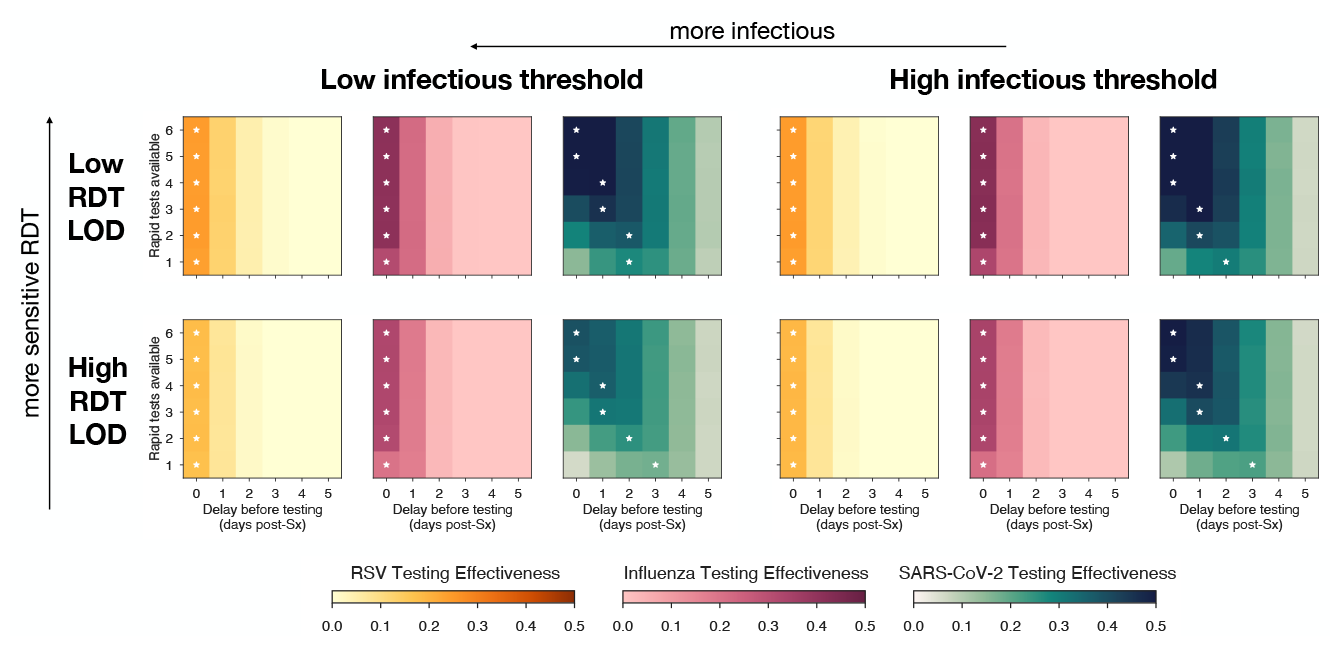
Sensitivity analysis of optimal timing of RDT usage after symptom onset. Testing effectiveness (TE) of rapid test (RDT), used *x* days after symptom (Sx) onset using *y* tests once per day is shown for RSV (orange), influenza A (pink), and SARS-CoV-2 omicron in experienced hosts (green). Darker colors represent higher TE as indicated. In each row, the testing strategy with highest TE is annotated with a white star. Each scenario is considered for a 0.5 log-fold increase or decrease in infectious threshold or RDT limit of detection (LOD), as labeled. In this sensitivity analysis, optimal test timing recommendations for RSV and influenza A are unaffected, and are shifted by at most 1d for SARS-CoV-2. Turnaround times: rapid tests, TAT = 0; RT-qPCR TAT = 2.

**Figure S7:**
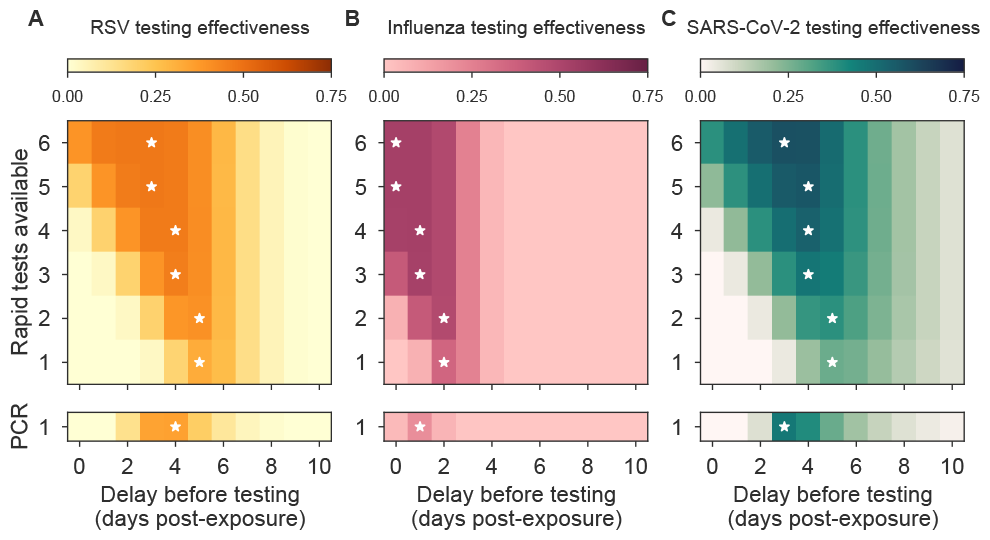
Optimal timing of testing after known exposure depends on the number of tests available. Testing effectiveness (TE) of rapid test (RDT) and RT-qPCR with 2 day turnaround time, used *x* days after exposure using *y* tests once per day is shown for RSV (orange), influenza A (pink), and SARS-CoV-2 omicron in experienced hosts (green). Darker colors represent higher TE as indicated. In each row, the testing strategy with highest TE is annotated with a white star. Turnaround times: rapid tests, TAT = 0; RT-qPCR TAT = 2. See Supplementary Table S1 for LODs and Figure 4 for TE estimates.

**Figure S8:**
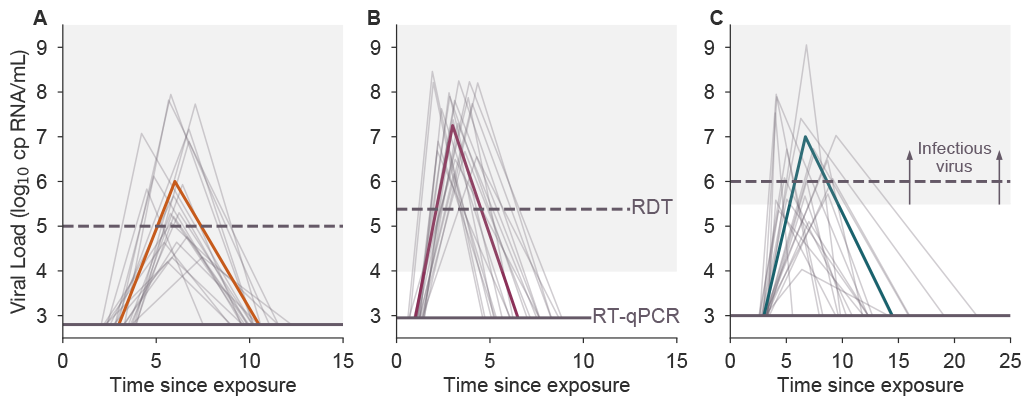
Examples of modeled viral kinetics. Stochastic realizations (gray lines; n=15) and a single characteristic realization (colored line) of the piecewise linear model of viral load kinetics for RSV (orange), influenza A (pink), and SARS-CoV-2 omicron/experienced (green). Horizontal lines show the pathogen-specific limit of detection of RT-qPCR (solid line) and rapid diagnostic tests (RDTs; dashed line). Infectious viral load is indicated by the gray shaded region.

**Table S1:** Summary of viral load, infectiousness, and testing parameters. For a description of how parameters were extracted from the cited sources, please see Materials and Methods. LOD, limit of detection; VL, viral load; Unif, uniform; ^*∗*^ bounded within [0.5, 10]; ^*†*^ bounded within [0.5, 25].

| Pathogen | Parameter | Value | Units | Source |
| --- | --- | --- | --- | --- |
| Influenza A | Latent period | Unif[0.5,1.5] | Days from exposure | [30,31] |
|  | Peak time | Unif[1,3] | Days from latent | [30–32] |
|  | Peak VL | Unif[6,8.5] | log<sub>10</sub> cp RNA/mL | [32] |
|  | Clearance time | Unif[2,5] | Days from VL peak | [30–32] |
|  | Infectious threshold | 4 | log<sub>10</sub> cp RNA/mL | [38] |
|  | High sensitivity LOD | 2.95 | log<sub>10</sub> cp RNA/mL | [32] |
|  | Low sensitivity LOD | 5.38 | log<sub>10</sub> cp RNA/mL | [73] |
|  | Percent symptomatic | 64 | Percent of infections | [30–32] |
|  | Symptom onset time | Unif[-2,0] | Days from VL peak | [38] |
|  | Failure rate | 5 | Percent of tests above LOD | See text |
| RSV | Latent period | Unif[2,4] | Days from exposure | [28,29] |
|  | Peak time | Unif[2,4] | Days from latent | [28] |
|  | Peak VL | Unif[4,8] | log<sub>10</sub> cp RNA/mL | [28,29] |
|  | Clearance time | Unif[3,6] | Days from VL peak | [28,29] |
|  | Infectious threshold | 2.8 | log<sub>10</sub> cp RNA/mL | [29] |
|  | High sensitivity LOD | 2.8 | log<sub>10</sub> cp RNA/mL | [28] |
|  | Low sensitivity LOD | 5 | log<sub>10</sub> cp RNA/mL | [74] |
|  | Percent symptomatic | 57 | Percent of infections | [39,71] |
|  | Symptom onset time | Unif[-1,1] | Days from VL peak | [40] |
|  | Failure rate | 5 | Percent of tests above LOD | See text |
| SARS-CoV-2<br>omicron strain<br>experienced host | Latent period | Unif[2.5,3.5] | Days from exposure | [13,33] |
|  | Peak time | Lognormal[1.053,0.688]* | Days from latent | [34] |
|  | Peak VL | Lognormal[1.876,0.181] | 40 - Cycle threshold | [34] |
|  | Clearance time | Lognormal[1.704,0.491]<sup>†</sup> | Days from VL peak | [34] |
|  | Infectious threshold | 5.5 | log<sub>10</sub> cp RNA/mL | [36,75–77] |
|  | High sensitivity LOD | 3 | log<sub>10</sub> cp RNA/mL | [33,34] |
|  | Low sensitivity LOD | 6 | log<sub>10</sub> cp RNA/mL | [45,46] |
|  | Percent symptomatic | 65 | Percent of infections | [34] |
|  | Symptom onset time | Unif[-5,-1] | Days from VL peak | [34,49,50] |
|  | Failure rate | 5 | Percent of tests above LOD | [45] |
| SARS-CoV-2<br>founder strain<br>naive host | Latent period | Unif[2.5,3.5] | Days from exposure | [13,33] |
|  | Peak time | Lognormal[0.873,0.788]* | Days from latent | [17] |
|  | Peak VL | Lognormal[1.999,0.199] | log<sub>10</sub> cp RNA/mL | [17] |
|  | Clearance time | Lognormal[1.953,0.611]<sup>†</sup> | Days from VL peak | [17] |
|  | Infectious threshold | 5.5 | log<sub>10</sub> cp RNA/mL | [36,75–77] |
|  | High sensitivity LOD | 3 | log<sub>10</sub> cp RNA/mL | [34] |
|  | Low sensitivity LOD | 5 | log<sub>10</sub> cp RNA/mL | [46] |
|  | Percent symptomatic | 65 | Percent of infections | [33,34] |
|  | Symptom onset time | Unif[0,3] | Days from VL peak | [33,68,69] |
|  | Failure rate | 5 | Percent of tests above LOD | See text |

## Supplementary Text: Imperfect Isolation Behaviors

To include imperfect post-diagnosis isolation behaviors, we consider a behavior change function *B*(*τ*), one’s relative infectiousness at time since diagnosis *τ*. For instance, perfect isolation upon diagnosis would be modeled as *B*(*τ*) = 0, while a one-week partial isolation might be modeled as *B*(*τ*) = 0.5 for 0 *≤ τ ≤* 7, and *B*(*τ*) = 1 for *τ >* 7. The individual reproduction number can thus be written as the sum of the total infectiousness before and after diagnosis,

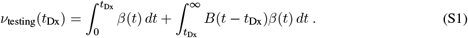

Simplifying yields

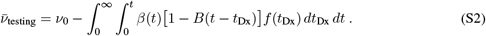

This equation offers a helpful term-by-term interpretation: diagnosis decreases total infectiousness from its baseline of *ν*_0_ by an amount that depends on (i) the probability distribution of diagnosis times *f* (*t*_Dx_) and (ii) the quality of isolation after said diagnosis *B*(*τ*), weighted by (iii) the infectiousness *β*(*t*) at the time of isolation and thereafter. Under a specified isolation behavior *B*(*τ*), TE is computed as

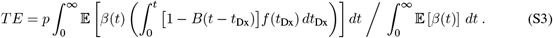

Thus, more complex post-diagnosis behaviors may be easily modeled, but such scenarios were not explored numerically in the main text.

## Supplementary Text: Test to Exit Strategies

Test-to-exit (TTE) is a strategy designed to maximize the effectiveness of post-diagnosis isolation while minimizing the number of days spent in isolation by requiring one or more negative test(s) before exiting isolation. While various formulations of TTE may exist, here we analyze a simple version in which individuals wait *w* days after receiving a diagnosis and then begin testing at a rate Ā tests per day, with a per-test failure rate of *ϕ* and a test turnaround time of TAT. We assume that compliance with TTE is *c* = 1 due to individuals’ expected desire to leave isolation.

Mirroring Eq. (S1), the expected infectiousness under a test-to-exit program, assuming perfect isolation between the time of diagnosis *t*_Dx_ and the time of isolation exit *t*_exit_, is given by

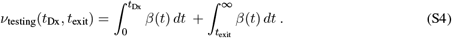

We define the PDF and CDF of *t*_exit_ as *g* and *G*, respectively, allowing us to rewrite the previous equation as

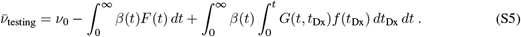

This expression takes on the interpretable form of total infectiousness in the absence of testing *ν*_0_, minus post-diagnosis infectiousness prevented due to perfect and indefinite isolation, plus any residual infectiousness realized by an exit from isolation.

To model post-isolation exit testing, we assume that one waits *w* days before testing at a rate *Ā* using a test with turnaround time TAT and failure rate *ϕ*. Under these conditions, *n*(*t*) = Ā (*t − t*_Dx_ *−* TAT *− w*) represents the expected cumulative number of scheduled tests with the potential to return an exit-inducing result by time *t*, with 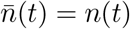 mod1. The distribution for *t*_exit_ is then given by the rather cumbersome

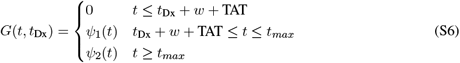

where the function

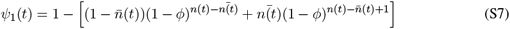

represents the cumulative probability that one receives a negative test due to a false negative (i.e., a test failure when above that test’s limit of detection), and where

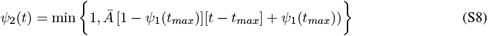

captures the rapid approach of *G* toward guaranteed exit from isolation after one is no longer detectable. The quantity *t*_*max*_ = max [*t*_*u*_ + TAT, *t*_Dx_ + *w* + TAT] represents the latest possible time at which a person testing to exit could receive a positive test. The corresponding PDF is

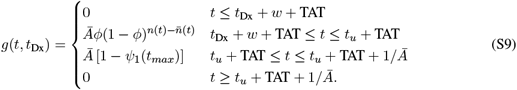

Under the above TTE assumptions, the typical number of days spent in isolation may be computed as the expected difference between the *t*_exit_ and *t*_Dx_ distributions. One may also update 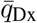 in computing test consumption (Materials and Methods) to include the additional tests consumed while testing to exit,

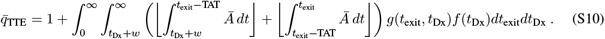

